# Estimating effective reproduction number using generation time versus serial interval, with application to COVID-19 in the Greater Toronto Area, Canada

**DOI:** 10.1101/2020.05.24.20109215

**Authors:** Jesse Knight, Sharmistha Mishra

## Abstract

**Background:** The effective reproduction number *R_e_*(*t*) is a critical measure of epidemic potential. *R_e_*(*t*) can be calculated in near real time using an incidence time series and the generation time distribution—the time between infection events in an infector-infectee pair. In calculating *R_e_*(*t*), the generation time distribution is often approximated by the serial interval distribution—the time between symptom onset in an infector-infectee pair. However, while generation time must be positive by definition, serial interval can be negative if transmission can occur before symptoms, such as in covid-19, rendering such an approximation improper in some contexts.

**Methods:** We developed a method to infer the generation time distribution from parametric definitions of the serial interval and incubation period distributions. We then compared estimates of *R_e_*(*t*) for covid-19 in the Greater Toronto Area of Canada using: negative-permitting versus non-negative serial interval distributions, versus the inferred generation time distribution.

**Results:** We estimated the generation time of covid-19 to be Gamma-distributed with mean 3.99 and standard deviation 2.96 days. Relative to the generation time distribution, non-negative serial interval distribution caused overestimation of *R_e_*(*t*) due to larger mean, while negative-permitting serial interval distribution caused underestimation of *R_e_*(*t*) due to larger variance.

**Implications:** Approximation of the generation time distribution of covid-19 with non-negative or negative-permitting serial interval distributions when calculating *R_e_*(*t*) may result in over or underestimation of transmission potential, respectively.

## 1 Introduction

The effective reproduction number *R_e_*(*t*) provides an instantaneous measure of transmission potential or the rate of spread of an epidemic. Cori et al. [1] provide a method to estimate *R_e_*(*t*) in quasi-real time based on only two inputs: an incidence time series,^1^ and the generation time distribution. The generation time is defined as the time between infection events in an infector-infectee pair.

When estimating *R_e_*(*t*) in previous epidemics [1–3] and in covid-19 [4–7], the generation time has been approximated by the serial interval. The serial interval is defined as the time between symptom onset in an infector-infectee pair. Unlike infection events, symptom onset is directly observable. This approximation is reasonable for infectious diseases where onset of infectiousness and symptoms is effectively simultaneous [1] such for SARS and Ebola [8, 9]. However, potential pre-symptomatic transmission of covid-19 [10, 11] renders approximation of generation time by serial interval problematic. Namely, while generation time is strictly positive, the serial interval can be negative, such as in 59 of 468 (12.6%) reported cases in [11], due to variability in the incubation period. As a result, covid-19 serial interval data have been used to fit both non-negative and negative-permitting distributions, yielding different estimates of *R_e_*(*t*) [11–13].

We compared estimates of *R_e_*(*t*) and their implications using each of the following approximations of the generation time distribution: (a) negative-permitting distribution fit to serial interval data; (b) non-negative distribution fit to serial interval data; and (c) inferred generation time distribution based on the incubation period and the serial interval distributions. We used parametric distributions described in the literature for covid-19, and reported cases from the Greater Toronto Area (gta), Canada.

## 2 Methods

First, we designed a simple method to recover the generation time distribution from parametric definitions of the incubation period and serial interval distributions, similar to work by Kuk et al. [14] and Britton et al. [15]. We then applied the method to covid-19 using published incubation period and serial interval distributions. Finally, we compared *R_e_*(*t*) estimated for the Greater Toronto Area (gta) region of Canada between March 08 and April 15, 2020 using the estimated generation time distribution versus negative permitting and non-negative serial interval distributions reported in the literature.

### 2.1 Estimating the Generation Time Distribution

Let *i* and *i* + 1 be the indices of an infector-infectee pair. Let *f_i_* and *f_i_*_+1_ be the respective times of infection, such that *g_i_* = [*f_i_*_+1_ − *f_i_*] ~ *G*(*τ*) is the generation time. Let *y_i_* be the time of symptom onset in case *i*, such that *h_i_* = [*y_i_* − *f_i_*] ~ *H*(*τ*) is the incubation period. Finally, let *s_i_* = [*y_i_*_+1_ − *y_i_*] ~ *S*(*τ*) be the serial interval. Figure 1 illustrates the variables and distributions graphically. We assume all distributions are independent, although previous work has shown that the generation time and incubation period may be correlated, for example, in the context of measles [16].

**Figure 1:**
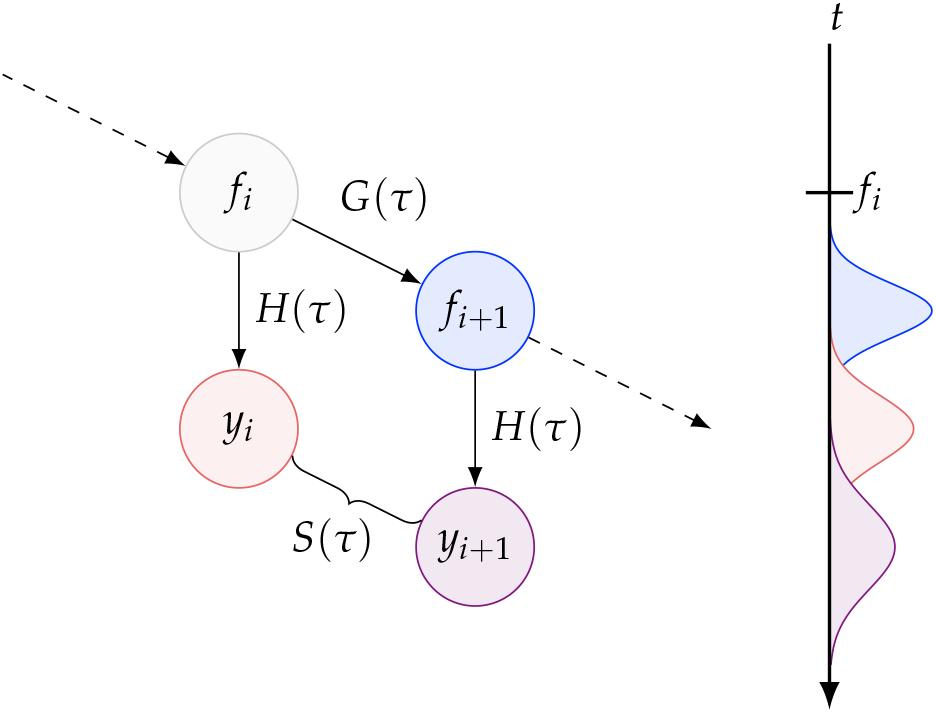
Random variables involved in the serial interval Notation — *i*: infector index; *i* + 1: infectee index; *f_i_*: time of infection; *y_i_*: time of symptom onset; *G*(*τ*): generation time distribution; *H*(*τ*): incubation time distribution; *S*(*τ*): serial interval distribution.

Rearranging, we have: *s_i_* = *g_i_* + *h_i_*_+1_ − *h_i_*. The probability distribution of the sum of independent random variables is the convolution of their respective distributions [17], where convolution *\** is defined as:

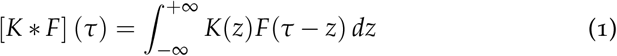

Thus *S*(*τ*) can be defined as:

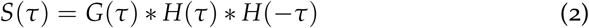

and *G*(*τ*) can be recovered using deconvolution *\*^−^*^1^ as:

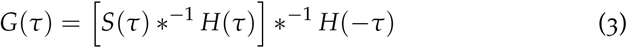

Some definitions of *S*(*τ*) and *H*(*τ*) may yield forms of *G*(*τ*) via deconvolution in (3) which are implausible or intractable. So, we defined a parametric form *Ĝ* (*τ | θ*), and found parameters *θ^*^* that minimized the Kullback-Leibler divergence between the observed *S*(*τ*) and *Ŝ*(*τ θ*) obtained via (2) using *Ĝ*(*τ | θ^*^*). It can be shown that such parameters *θ^*^* provide the maximum likelihood estimate (mle) of *S*(*τ*) under *Ĝ*(*τ | θ*).

### 2.2 Application

#### 2.2.1 Generation Time

We identified several parameterizations of the covid-19 incubation period and serial interval following the review by Park et al. [18] (Table A.1).^2^ For our analysis, we used the negative-permitting serial interval from [11] (*N* = 468):

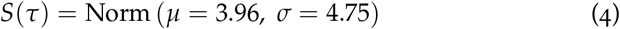

and the incubation period from [19] (*N* = 181):

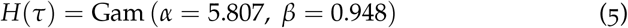

We assumed a Gamma parametric form for the generation time distribution *Ĝ*(*τ | θ*), with *θ* = [*α* (shape), *β* (scale)], for consistency with downstream assumptions used in calculating *R_e_*(*t*). We then minimized the KullbackLeibler divergence between *S*(*τ*) and *Ŝ*(*τ|θ*) using the Nelder-Mead optimization method in the optimization R package^3^ to obtain the mle generation time distribution parameters *θ^*^*.

### 2.2.2 Effective Reproduction Number

In the model described by Cori et al. [1], the incidence *I* at time *t* is given by the integral over all previous infections, multiplied by their respective infectivity *ω* at time *τ* since infection, collectively multiplied by the effective reproduction number *R_e_* at time *t*. This model yields the following definition of *R_e_*(*t*):

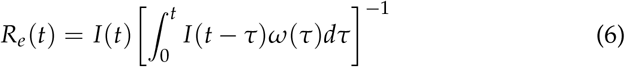

The infectivity profile *ω*(*τ*) is equivalent to the generation time distribution *G*(*τ*). Given *I*(*t*) and *G*(*τ*), probabilistic estimates of *R_e_*(*t*) can then be resolved in a Bayesian framework, as implemented in the EpiEstim R package.^4^

In order to quantify *R_e_*(*t*) of covid-19 in gta, Canada, we used reported cases in the region with episode dates between March 09 and May 04 2020 as the incidence time series *I*(*t*).^5^ We smoothed *I*(*t*) using a Gaussian kernel with *σ* = 1 day to reflect uncertainty in reporting delay. We then compared estimates of *R_e_*(*t*) using the mle generation time distribution versus serial interval distributions reported in the literature, including negative-permitting (Normal [11]), and non-negative (Gamma [12], Log-Normal [20]) distributions.

## 3 Results

Figure 2 shows the serial interval and incubation period distributions from [11] and [19] respectively, and the generation time distribution estimated via the proposed method. The mle parameters of *Ĝ*(*τ | θ*) were: shape *α* = 1.813 and scale *β* = 2.199, yielding *Ŝ*(*τ | θ^*^*) that reasonably approximated the target *S*(*τ*).

**Figure 2:**
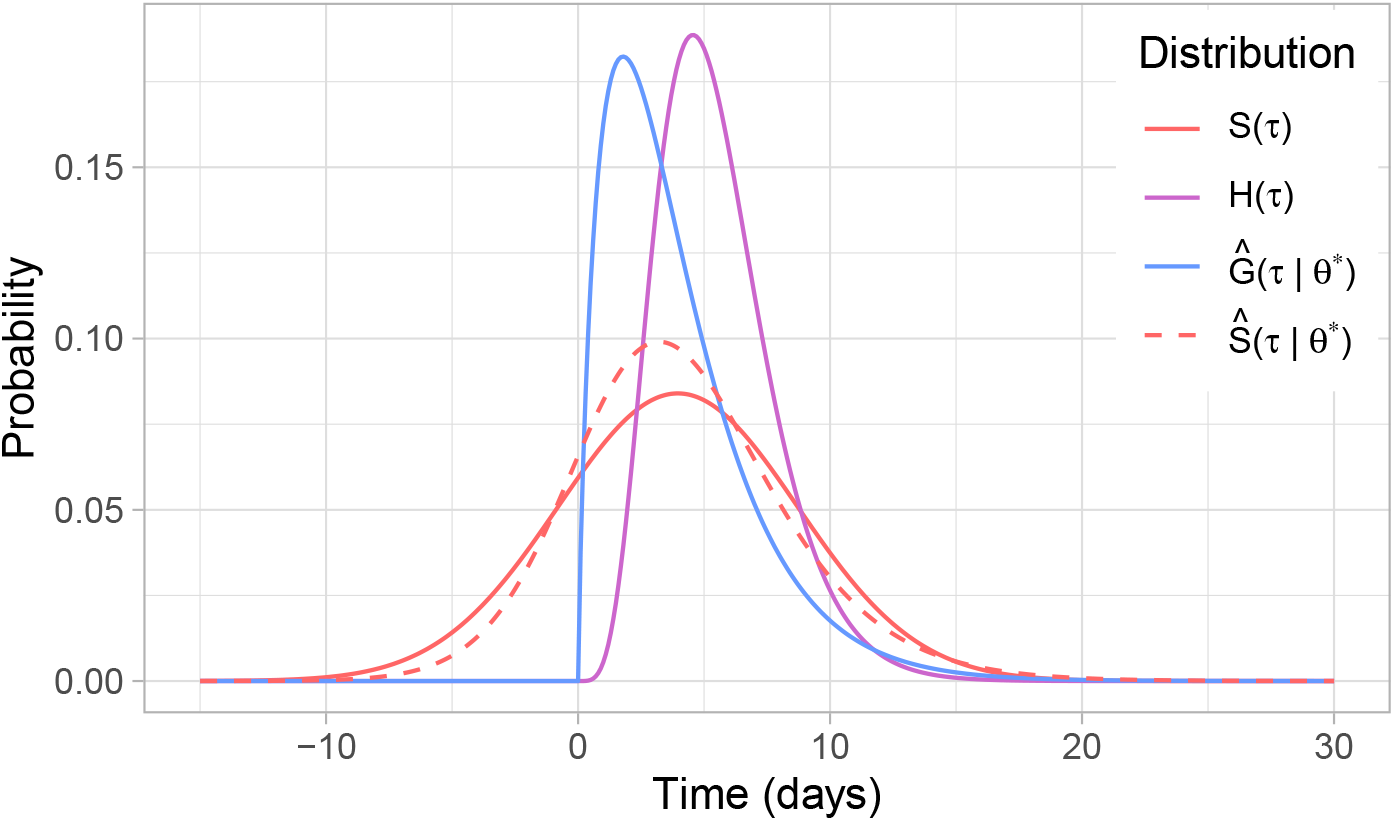
Recovered generation time distribution *Ĝ*(*τ | θ^*^*) based on mle approximation of the serial interval distribution *S*(*τ*) by *Ŝ*(*τ | θ^*^*) and the incubation period distribution *H*(*τ*).

Comparing the estimated generation time to published serial interval distributions (Figure A.1) showed the following. The mean generation time of 3.99 was similar to the mean serial interval of 3.96 based on the negative-permitting distribution [11], but shorter than mean serial interval based on non-negative distributions, such as 5.1 in [12] and 4.7 in [20]. The sd of the generation time distribution was smaller at 2.96 than the sd of the negative permitting serial interval at 4.75 [11], but similar to the sd of the non-negative serial interval distributions, including 2.7 in [12] and 2.9 in [20].

Figure 3 shows *R_e_*(*t*) for covid-19 in gta, Canada based on reported cases and estimated using the generation time distribution versus selected serial interval distributions reported in the literature.^6^ The *R_e_*(*t*) based on non-negative serial interval distribution was higher versus *R_e_*(*t*) using the estimated generation time distribution (e.g. using [12]: 2.33 vs 1.85 on March 16; and 1.32 vs 1.24 on April 13). Higher *R_e_*(*t*) can be attributed to longer mean serial interval under non-negative distributions versus the estimated mean generation time, since inferred *R_e_*(*t*) must be higher to compensate for longer delay between infections.^7^ By contrast, the *R_e_*(*t*) estimated using a negative-permitting serial interval distribution was the smallest of all three approaches (e.g. using [11]: 1.68 on March 16; and 1.22 on April 13). In this case, lower *R_e_*(*t*) can be attributed to increased variance in the negative-permitting serial interval distribution versus in the generation time distribution, as shown in [15].

**Figure 3:**
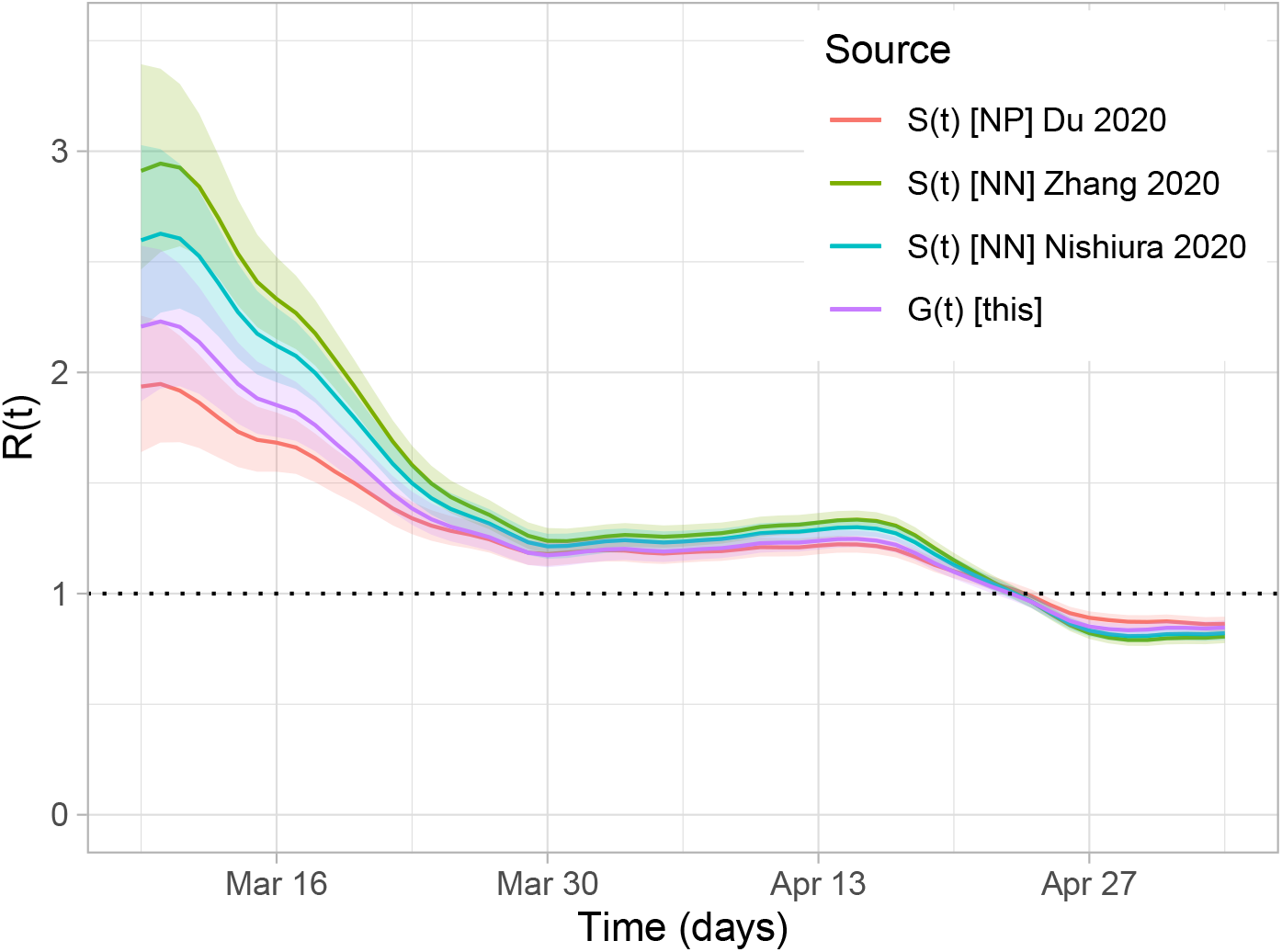
*R_e_*(*t*) of covid-19 in gta using serial interval versus generation time Notation — *S*(*τ*): serial interval; *G*(*τ*): generation time; [NP]: negative-permitting; [NN]: non-negative. See Figure A.2 for zoom-in of later dates.

## 4 Discussion

We have demonstrated how to estimate a non-negative generation time distribution based on negative serial interval and non-negative incubation period parametric distributions. We showed that estimates of *R_e_*(*t*) will vary depending on the approach used to approximate/estimate the generation time distribution. Specifically, relative to the estimated generation time distribution, non-negative and negative-permitting serial interval distributions may over and underestimate *R_e_*(*t*), respectively.

Our estimated generation time for covid-19 was Gamma-distributed with mean 3.99 and sd 2.96, similar to results by Ganyani et al. [13] (Table A.1). However in [13] person-level serial interval data on infector-infectee pairs are required for a joint Bayesian model of generation time, incubation period, and serial interval, characterized via mcmc sampling. In our approach, we used parametric distributions as inputs because we did not have access to person-level and paired data. As such, our approach could also use pooled estimates via meta-analyses of serial interval and incubation period as parameter inputs.

In several recent works [12, 21–23], non-negative serial interval distributions have been used as an approximation of the generation time distribution when estimating *R_e_*(*t*) for covid-19.^8^ We found that such an approximation may result in overestimation of *R_e_*(*t*), and thus overestimation of covid-19 transmission potential. The finding that *R_e_*(*t*) may be overestimated when using (non-negative) serial interval versus generation time seemingly contradicts the conclusion of Britton et al. [15], but can be explained as follows. In our study of covid-19, the mean serial interval under non-negative distributions [12, 20] was longer than the mean generation time estimated using negative-permitting serial interval [11]. If time between infections is assumed to be longer, then we would *overestimate R_e_*(*t*) to fit the same incidence. By contrast, Britton et al. [15] modelled both serial interval and generation time as Gamma-distributed (non-negative), resulting in equal means, but different variances. They then showed how increased variance in the serial interval distribution can cause *underestimation* of *R_e_*(*t*), relative to the generation time distribution. In fact, we also find underestimation of *R_e_*(*t*) when comparing negative-permitting serial interval to the generation time distribution, which had equal means.

Our approach to estimating generation time had three notable limitations. First, like similar works [13, 14], we assumed that generation time and incubation period were independent, although Klinkenberg et al. [16] showed that correlation between the two exist in infections such as measles. Second, as noted above, our approach did not use person-level serial interval or incubation period data such as in [13] and [16], possibly resulting in compounding errors from parametric approximation of both input (serial interval, incubation period) and output (generation time) distributions. However, depending on the availability and reliability of person-level data, our approach could in some cases be favourable. Finally, we did not perform uncertainty analysis using the reported confidence intervals for the serial interval and incubation period distribution parameters. Future work could overcome this limitation by exploring joint estimation of generation time, serial interval, and *R_e_*(*t*) within the Bayesian framework defined in [1].

## Data Availability

Reported COVID-19 cases were obtained from the Public Health Ontario (PHO) integrated Public Health Information System (iPHIS), via the Ontario COVID-19 Modelling Consensus Table. Such data are not currently publicly available.
Analysis code for inferring the generation time distribution and for estimating Re(t) is publicly available on GitHub, including parametric definitions of serial interval, incubation period, and generation time distributions from the literature.

https://github.com/mishra-lab/covid19-Re

## Additional Information

## Acknowledgements

We thank Kristy Yiu, Linwei Wang, Huiting Ma, and David Landsman (MAP Centre for Urban Health Solutions, Unity Health Toronto) for their help cleaning and validating covid-19 data used in these analyses. We thank David Champredon (University of Western Ontario) for helpful comments on a preliminary draft of the report and work.

Reported covid-19 cases were obtained from the Public Health Ontario (PHO) integrated Public Health Information System (iPHIS), via the Ontario covid-19 Modelling Consensus Table, and with approval from the University of Toronto Health Sciences REB (protocol No. 39253).

## Contributions

*Jesse Knight*: Conceptualization, Methodology, Formal Analysis, Investigation, Software, Visualization, Writing – drafting & editing; *Sharmistha Mishra*: Conceptualization, Investigation, Writing – review & editing, Resources, Funding acquisition.

## A Data & Code

All code and results for this project is available online at: https://github.com/mishra-lab/covid19-Re

Table A.1 summarizes the reported parametric covid-19 distributions, and Figure A.1 illustrates the serial interval and generation time distributions explored for estimating *R_e_*(*t*). Figure A.2 provides a zoom-in of Figure 3 after March 30.

**Table A.1:** Summary of reported parametric covid-19 distributions

|  | Ref | N | Distribution | Mean | SD |
| --- | --- | --- | --- | --- | --- |
| $S(\tau)$ | Du et al. [11] | 468 | Norm ($\mu = 3.96, \sigma = 4.75$) | 3.96 | 4.75 |
| | Zhang et al. [12] | 35 | Gam ($\alpha = 3.619, \beta = 1.416$) | 5.1 | 2.7 |
| | Nishiura et al. [20] | 28 | LogN ($\mu = 4.7, \sigma = 2.9$) | 4.7 | 2.9 |
| | Zhao et al. [25] | 21 | Gam ($\alpha = 2.151, \beta = 2.045$) | 4.4 | 3.0 |
| | Li et al. [24] | 6 | Gam ($\alpha = 4.866, \beta = 1.541$) | 7.5 | 3.4 |
| | Tindale et al. [26] | 4 | Norm ($\mu = 4.22, \sigma = 0.4$) | 4.22 | 0.4 |
| | Tindale et al. [26] | 4 | Norm ($\mu = 4.56, \sigma = 0.95$) | 4.56 | 0.95 |
| $H(\tau)$ | Lauer et al. [19] | 181 | Gam ($\alpha = 5.807, \sigma = 0.948$) | 5.51 | 2.28 |
| | Tindale et al. [26] | 135 | Weib ($\alpha = 2.25, \beta = 10.15$) | 8.99 | 4.23 |
| | Tindale et al. [26] | 93 | Weib ($\alpha = 1.88, \beta = 7.97$) | 7.07 | 3.91 |
| | Linton et al. [27] | 52 | LogN ($\mu = 5.6, \sigma = 2.8$) | 5.6 | 2.8 |
| | Backer et al. [28] | 88 | Weib ($\alpha = 3.038, \beta = 7.163$) | 6.4 | 2.3 |
| | Li et al. [24] | 10 | LogN ($\mu = 5.2, \sigma = 3.91$) | 5.2 | 3.91 |
| $G(\tau)$ | this | — | Gam ($\alpha = 1.813, \beta = 2.199$) | 3.99 | 2.96 |
| | Ganyani et al. [13] | * | Gam ($\alpha = 9.140, \beta = 0.569$) | 5.20 | 1.72 |
| | Ganyani et al. [13] | * | Gam ($\alpha = 6.843, \beta = 0.577$) | 3.95 | 1.51 |

Notation — $N$ : sample size; \*: indeterminate; $S(\tau)$ : serial interval; $H(\tau)$ : incubation time; $G(\tau)$ : generation time; $\mu$ : mean; $\sigma$ : std dev (sd); $\alpha$ : shape; $\beta$ : scale. Notes — some parameters were back-calculated based on reported distribution statistics like mean, sd, and quantiles; LogN mean and sd are in linear, not log scale.

**Figure A.1:**
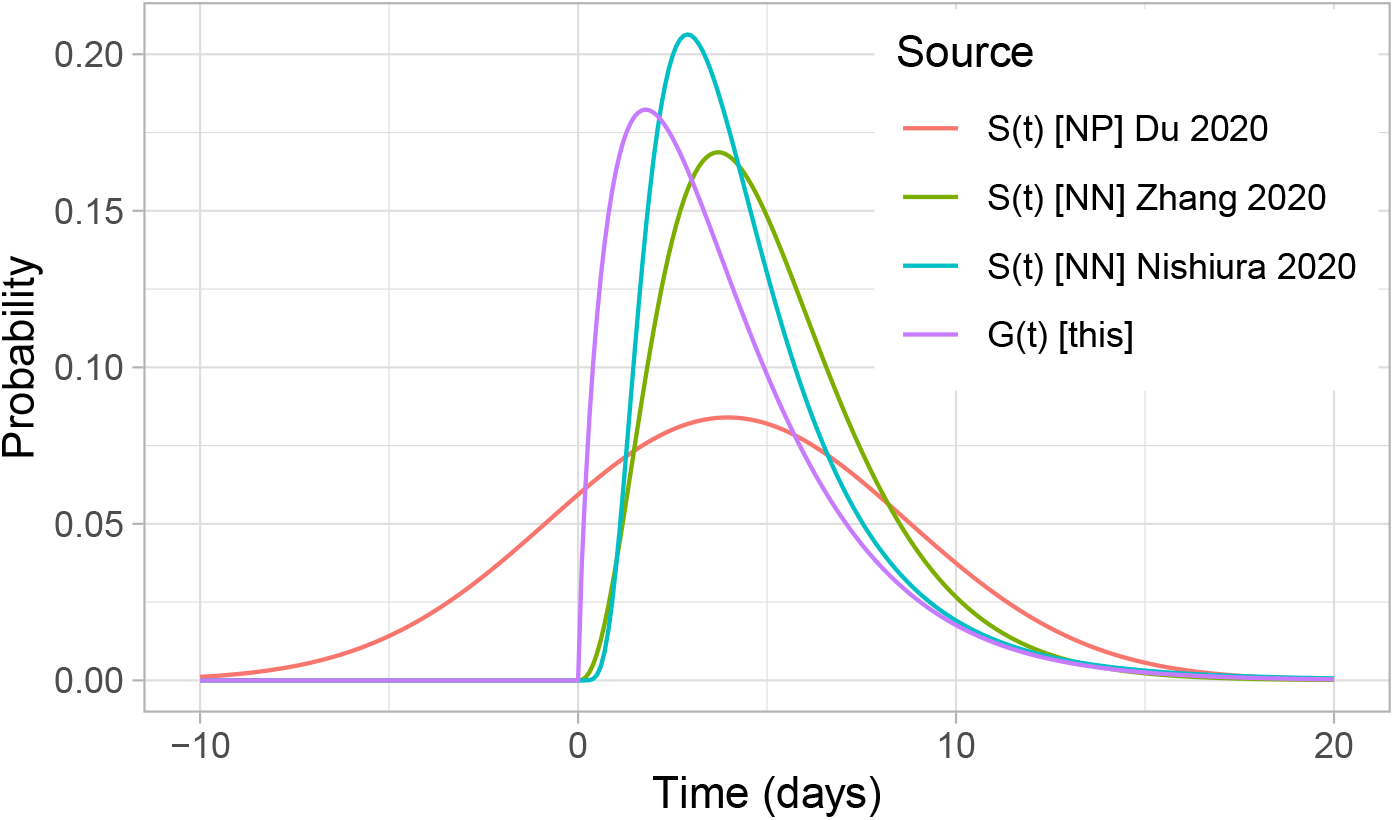
Illustration of reported serial interval and generation time distributions used for calculating *R_e_*(*t*) in covid-19

**Figure A.2:**
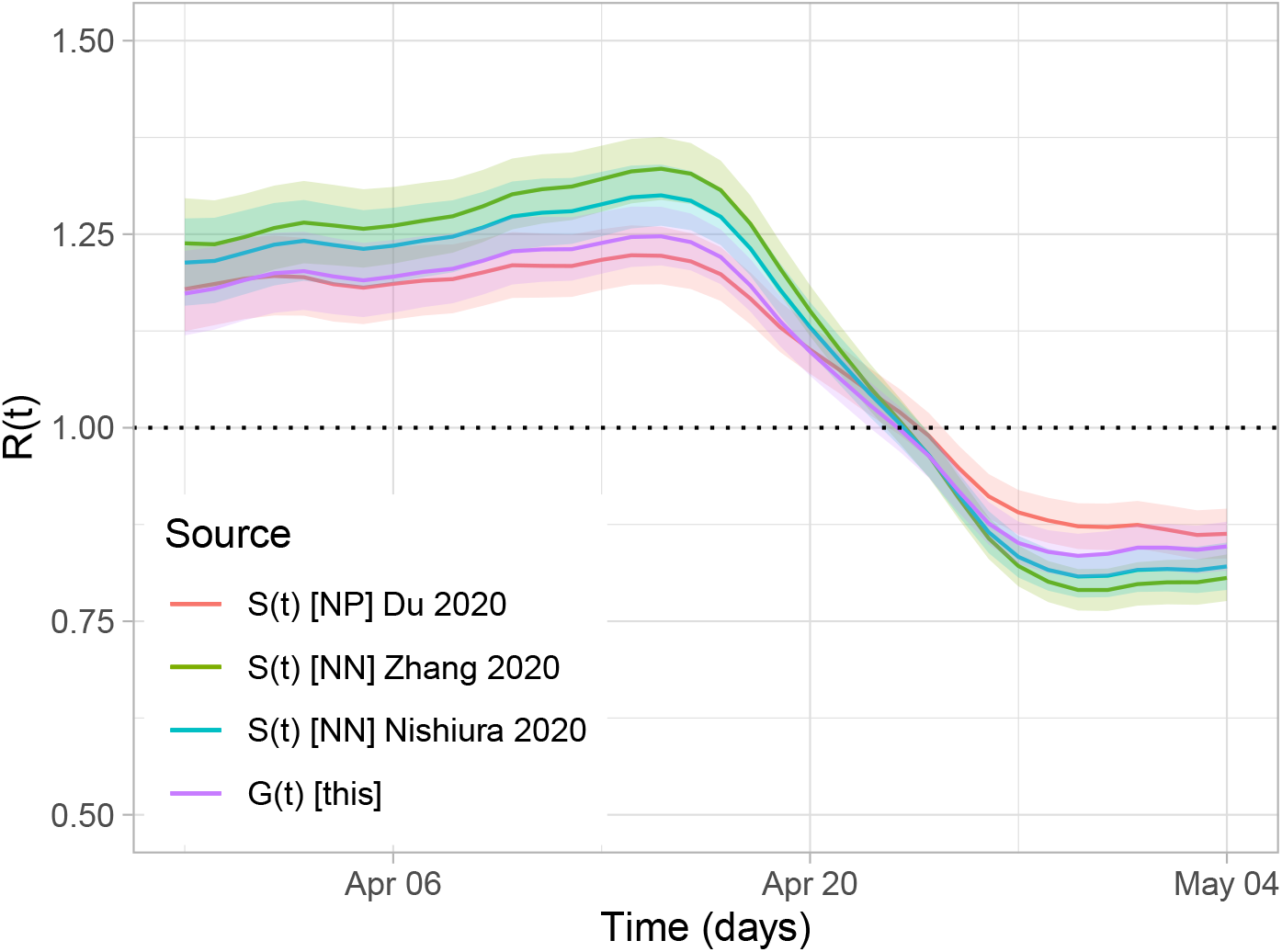
*R_e_*(*t*) of covid-19 in gta using serial interval versus generation time (zoom of March 30 to May 4) Notation — *S*(*τ*): serial interval; *G*(*τ*): generation time; [NP]: negative-permitting; [NN]: non-negative.

## Footnotes

1 Incidence may be further stratified by imported versus locally generated cases to quantify local transmission dynamics.

2 Some studies did not provide enough information to define a parametric form (e.g. only reported the mean).

3 https://cran.r-project.org/web/packages/optimization

4 https://cran.r-project.org/web/packages/EpiEstim

5 We did not stratify incidence by “imported” vs “local” transmission events, in order to quantify overall epidemic growth. The relative influence of different *G*(*τ*) approximations on *R_e_*(*t*) should not be affected by this lack of stratification.

6 Generation time and serial interval distributions are also illustrated in Figure A.1.

7 Relative differences in *R_e_*(*t*) between input distributions are “flipped” after *R_e_* < 1, since a longer delay between infections has opposite implications for *R_e_*(*t*) in the context of a shrinking versus growing epidemic.

8 In early characterizations of covid-19 [12, 20, 21, 24, 25], negative serial interval (and presymptomatic transmission) was either unobserved or considered implausible.

